# Determinants of motivated behavior are linked to fatigue and its perturbation by SARS-CoV-2 vaccination

**DOI:** 10.1101/2022.04.23.22274186

**Authors:** David S. Stolz, Finn Luebber, Tanja Lange, Stefan Borgwardt, Malte Ziemann, Gabriela Riemekasten, Jan Rupp, Laura Müller-Pinzler, Frieder M. Paulus, Sören Krach

## Abstract

**Background:** Fatigue has an adaptive function and serves as a temporary signal to rest and save energy often in response to immune activation. It may, however, also persist in a pathological condition incurring significant burden. While subjective symptoms and scientific consensus indicate that both physical and mental determinants of motivated behavior are affected in fatigue, the underlying processes are rarely examined using objective, task-based indicators.

**Methods:** In three consecutive studies, including validation (*N* = 48) and reliability assessments (*N* = 27), we use an experimental task to jointly objectify reward learning and effort execution as two determinants of behavioral motivation. In addition, we tested how fatigue and its acute perturbation in response to immune activation after SARS-CoV-2 vaccination are linked to these task-based indicators of motivation in a longitudinal cross-over design (*N* = 55).

**Results:** The validation study showed the utility of the experimental task for simultaneously assessing learning, effort exertion, and its regulation based on subjective confidence. The reliability assessment over a one-week period indicated that symptoms of fatigue and task behavior are highly reliable and that repetition effects have little impact on motivated behavior. Finally, in the vaccination trial, we found significant links between fatigue and task behavior. Baseline levels of fatigue predicted how effort is gauged in dependence of current confidence about reward outcomes, and state perturbations of fatigue in the context of the SARS-CoV-2 vaccination reduced confidence during learning. Importantly, task success was significantly lower in subjects who reported high fatigue at baseline and who additionally experienced stronger increase in fatigue in response to vaccination.

**Discussion:** Our results demonstrate that the experimental task allows to jointly assess determinants of motivated behavior, and to link its constituent processes to subjective fatigue. This suggests that our understanding of fatigue and its perturbation due to acute immune activation can benefit from objective, task-based indicators of the underlying motivational mechanisms. Future studies could build on these findings to further deepen the understanding of neurobehavioral mechanisms underlying fatigue in the context of immune activation.

## Introduction

Since the onset of the COVID-19 pandemic, public and scientific interest in understanding fatigue as a short-term, but also potential long-term consequence of an infection with SARS-CoV-2 and beyond has significantly increased (Islam et al., 2020; Komaroff & Bateman, 2020; Townsend et al., 2020; Wostyn, 2021). Fatigue, first and foremost, is an adaptive response to perceived exhaustion of the body and serves as a signal to rest and save energy (Keyser, 2010; Lacourt et al., 2018). However, the experience of fatigue has significant consequences for the motivation and execution of even light everyday life activities, with enduring states resulting in pathological fatigue associated with reduced physical and mental health (Afari & Buchwald, 2003). While the study of fatigue has made significant advances recently, it is yet unclear how fatigue due to immune activation relates to objectively quantifiable processes underlying motivated behavior. Filling this gap is important to more clearly delineate and integrate the immunological (Dantzer et al., 2014; Felger & Treadway, 2017), neural (Müller & Apps, 2019), computational (Stephan et al., 2016), and psychological (Hockey, 2010) mechanisms in the etiology of fatigue.

Today, fatigue is understood as an inherently multidimensional construct with at least physical and mental components affecting motivated behavior (Karshikoff et al., 2017; Lasselin, 2021), that can be assessed using subjective, questionnaire-based measures (Smets et al., 1995) and objective, task-based indicators (Müller et al., 2021; van der Schaaf et al., 2018). Here, the physical dimension of fatigue refers to bodily exhaustion and the perceived costs of the execution of physical tasks that require any kind of bodily effort (Müller & Apps, 2019). Physical task execution and effort expenditure are usually assessed with devices such as hand dynamometers or ergometers which have been used with various intensities and durations (Jäkel et al., 2021; Müller et al., 2021; van der Schaaf et al., 2018). The mental component of fatigue refers to perceived exhaustion while performing cognitive tasks, such as working memory (Lange et al., 2005), sustained attention (Schwid et al., 2003), or executive control (van der Linden et al., 2003). In the context of the enduring symptoms of a SARS-CoV-2 infection, often termed “Long-COVID” (Davis et al., 2021; World Health Organization, 2021), these have also been summarized with the subjective experience of “brain-fog” (Hugon et al., 2022). However, although the multidimensionality of fatigue is widely acknowledged, the interdependence of its components has so far been largely neglected, which limits the understanding of fatigue as a complex phenomenon with potentially diverse etiology.

Theoretical and experimental research aiming to explain how humans motivate behavior and succeed in their environments align with the multidimensional perspective of fatigue and acknowledge the importance of the different components and their interaction. Classical theories emphasize that besides showing effort and being able to correctly execute a certain behavior (reflecting the physical component of fatigue), it is necessary to learn which actions or how much effort is necessary to reach a desired goal (mental component) (Ajzen, 2002; Bandura, 1977). Precisely, learning the contingencies between actions and their rewarding outcomes leads to increased confidence about which actions result in desired states (Kepecs & Mainen, 2012; Yeung & Summerfield, 2012) and the experience of control over the environment (Huys & Dayan, 2009). The experience of control can be understood as a subjective meta-belief closely associated with positive affect and increased motivational states (Stolz et al., 2020) as well as health and mental well-being (Stephan et al., 2016). Yet, although the two components underlying motivated behavior – effort expenditure and reward learning – have been objectively quantified with experimental tasks (Harrison et al., 2016; Hauser et al., 2017; van der Schaaf et al., 2018), the influence of fatigue on their dynamic interplay has so far been largely neglected. This becomes relevant for understanding motivational consequences of fatigue in its various manifestations in the spectrum from an adaptive response (Keyser, 2010; Müller & Apps, 2019), to enduring pathological conditions as seen in various diseases (Chaudhuri & Behan, 2004; Stephan et al., 2016). Studies that consider only single aspects of fatigue or independently assess physical and mental components of motivated behavior might set unnecessary boundaries in advancing the understanding of fatigue.

When studying the effects of fatigue on motivated behavior, it is relevant to consider the different timescales in which fatigue varies within and between individuals (Boolani & Manierre, 2019). Changes in fatigue can reflect slow frequency dynamics and more enduring progressions towards chronic states as well as acute perturbations that are caused by temporarily circumscribed events. Relatively enduring differences in fatigue have been associated with life-style variables (Boolani & Manierre, 2019), personality traits (e.g. serenity, neuroticism; De Vries & Van Heck, 2002; Merkelbach et al., 2003), skills (e.g. coping behavior; Ax et al., 2001), or chronic health conditions (Induruwa et al., 2012; Nikolaus et al., 2013), and are linked to rather stable environmental factors (e.g. socio-economic status; Engberg et al., 2017). In contrast, more short-term changes in fatigue are linked foremost to temporally constrained events such as acute viral infections (Carrat et al., 2008), or environmental factors such as temporary stress (Doerr et al., 2015). As previous research has shown, the effects of acute events on behavioral regulation can differ depending on whether or not they are accompanied by more or less severe baseline conditions (Haeffel et al., 2008; Swanepoel et al., 2020), similar to vulnerability models of diseases (Ingram & Luxton, 2005).

In the present study, we introduce an experimental task to jointly objectify components of motivated behavior, i.e. effort expenditure and reward learning, in the context of vaccination against SARS-CoV-2. In a cross-over, repeated-measures design, we use the vaccination as an immune-system activation model to bridge the literature of fatigue and motivated behavior. In doing so, we aim to identify those aspects of behavioral regulation that have the potential to unveil novel aspects related to the burden and reduced well-being associated with fatigue. In three consecutive studies (overall *N*=130), we first validate our task (*n*=48) and show that effort expenditure increases with learning and confidence about the circumstances under which own actions result in desired outcomes. In study 2, involving a separate sample (*n*=27; repeated-measures in a seven-day interval), we show that parameters of motivated behavior can be reliably assessed with the experimental task and that these measures reflect rather stable individual differences. In study 3, involving a third independent sample (*n*=55; repeated-measures, cross-over design), we demonstrate that SARS-CoV-2 vaccination can serve as a temporarily limited immune-system activation model to study state changes of fatigue and its effects on behavior. We show that greater vaccination-induced state changes of fatigue are related to reduced confidence while learning when to express effort. Further, greater baseline fatigue predicts the dependency between effort expenditure and confidence. In line with vulnerability-stress models of disease (Ingram & Luxton, 2005), task performance decreases in participants with high baseline levels of fatigue who additionally show high state changes due to the vaccination.

## Results

### Learning of effort efficacy

In experiment 1, subjects (*N* = 48) performed the Learn/Effort-Task (**Fig. 1**). In brief, on each of 200 trials they chose between two colored buttons (yellow or blue). After this choice, subjects pressed a hand-dynamometer knowing that depending on whether the choice was correct or not, they would be either rewarded or penalized with up to 100 points (depending on the percentage of their maximum voluntary contraction, MVC). For example, if a subject chose the correct button and then exerted 78% of their MVC, 78 points were gained. If, however, the incorrect button had been chosen, 78 points were subtracted (see Methods for details). Thus, to maximize their feedback, subjects had to exert more effort if it was likely to be rewarded, and reduce their effort if it was likely to be penalized. Therefore, subjects needed to learn when choosing the blue or yellow button was likely to turn their effort into a reward (or punishment) and use this knowledge to regulate their effort expenditure on every trial.

**Figure 1.**
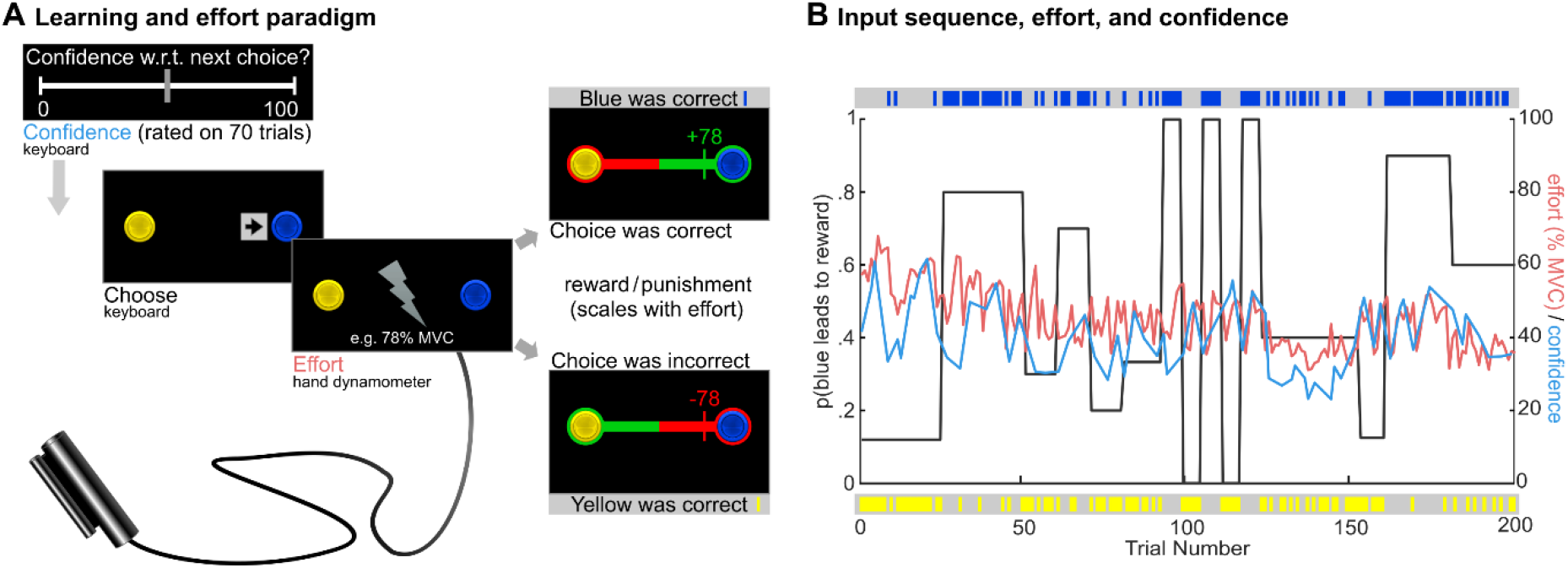
Overview of the experimental paradigm used in all three experiments. **A** Schematic depiction of steps in a single trial. **B** Depiction of the input sequence defining on which trials choosing the blue button would lead to reward, and yellow to punishment, and vice versa. Blue and yellow dashes on top and bottom indicate correct button for every single trial. Black line depicts probability with which blue is correct for different phases of the task. Light blue and red lines depict trial-by-trial effort (in % maximum voluntary contraction, MVC) and confidence ratings, respectively, averaged across all participants in experiment 1

Overall, subjects selected the correct button on 59.12% of trials (*SD*=3.48), illustrating that they learned about the probabilistic structure of the task (significant difference to chance level; *t*(47)=18.15, *p*<.001, *d*=2.62, 95%-CI=[2.02;3.22]; **Fig. 2**). Importantly, the confidence subjects experienced during learning predicted subsequent effort expenditure, as indicated by a significant positive correlation (mean *r*=.43, *SD*= .20; *t*(47)=10.58, *p*<.001). Across all trials, subjects received an average of 9.73 points (*SD*=4.41), which was significantly more than would have been expected if subjects had shown equal amounts of effort on correct and incorrect trials (*t*(47)=8.67, *p*<.001, *d*=1.25, 95%-CI=[0.87;1.63]; see Methods). Thus, when having the opportunity to freely decide whether to invest physical effort or not, subjects were successful in exploiting the acquired knowledge on the probabilistic structure of their environment. Voluntary effort expenditure increased with greater confidence about the opportunities to maximize rewards and decreased with lower confidence so as to avoid punishment. Here, in line with models about learning and behavioral control (Huys & Dayan, 2009; Stephan et al., 2016), the meta-belief of confidence reflects the expected probability of choosing correctly and is a central component of motivated behavior (Kepecs & Mainen, 2012; Navajas et al., 2017). Comparable belief-based regulation of effortful behavior has been shown in animal models (Kepecs et al., 2008) and studies involving humans (Hauser et al., 2017). Importantly, when organisms fail to establish a belief that their actions are efficacious, this entails reduced effort expenditure and eventually fatigue (Stephan et al., 2016), as has been prominently proposed in the concept of learned helplessness (Maier & Seligman, 1976).

**Figure 2.**
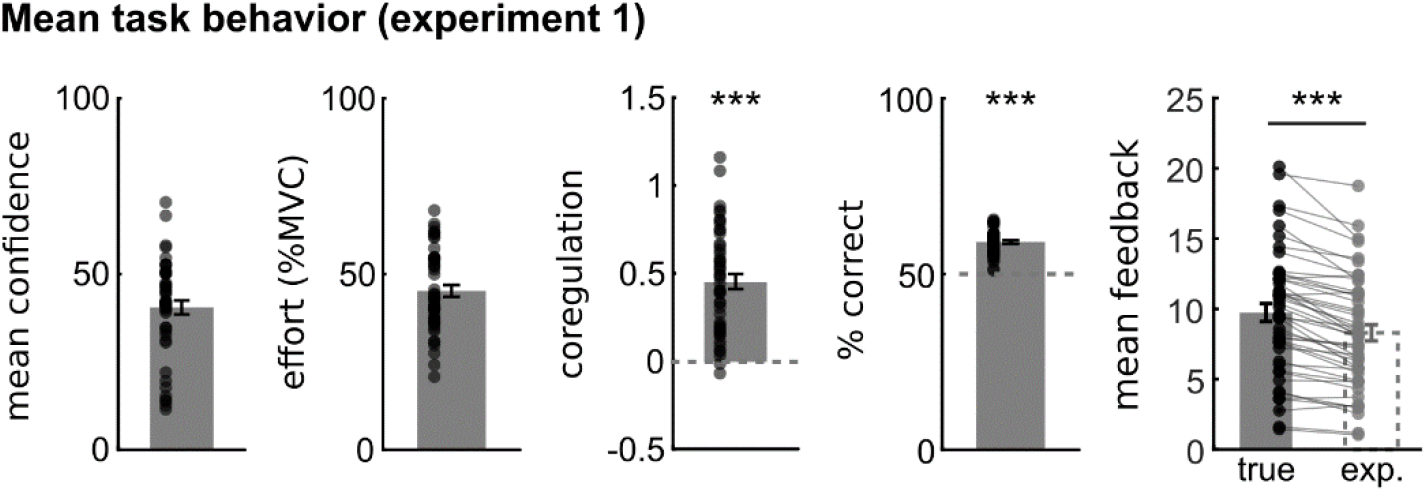
Results from experiment 1. Dashed lines in coregulation and % correct plots indicate comparison values for one-sample t-tests. Dashed boxplot with light grey dots displays hypothetical feedback that would be expected if subjects did not show different levels of effort on correct and incorrect trials (see main text for details). Error bars display +/-1 standard error of the mean. *** *p*<.001.

### Stability of fatigue and motivated behavior

Experiment 2 examined the stability of task parameters as well as fatigue scores. Subjects (*N*=27) were invited on two timepoints, separated by a median of 7.99 days (*IQR*=7.91), and performed the Learn/Effort-Task (two parallel versions) on each occasion. The task was identical to experiment 1 on the first timepoint. On the second timepoint the only two differences were that blue and yellow were replaced by purple and orange, and the contingency between the sequence of correct buttons and location on the screen (left vs. right) was inverted. However, the input sequence itself remained identical. In addition to the task, subjects filled in the 20-item Multidimensional Fatigue Inventory (MFI-20; Smets et al., 1995) on both test days, revealing high temporal stability of this measure (*r*=.87, *p*<.001, 95%-*CI*=[.74; .94]; **Fig. 3A**). This implies that individual differences in fatigue, if not otherwise perturbed by significant short-term events, reflect rather enduring individual characteristics that have previously been associated with life-style variables, personality traits, or chronic health conditions (e.g. Boolani & Manierre, 2019; De Vries & Van Heck, 2002; Induruwa et al., 2012). For the Learn/Effort-Task, all findings from experiment 1 replicated (**Fig. 3B**). On both days, the percentage of correct choices was better than chance (day 1: *M*=57.91%, *SD*=4.03%, *t*(26)=10.20, *p*<.001, *d*=1.96, 95%-*CI*=[1.31; 2.60]; day 2: *M*=59.07%, *SD*=4.23%, *t*(26)=11.15, *p*<.001, *d*=2.15, 95%-*CI*=[1.45; 2.83]), and as before, confidence and effort showed robust coregulation (day 1: mean *r*=.25, *SD*=0.30, *t*(26)=4.19, *p*<.001; day 2: mean *r*=.33, *SD*=.24, *t*(26)=6.47, *p*<.001). Again, this resulted in higher feedback than would have been expected if subjects had shown equal effort on correct and incorrect trials (day 1: *M*=7.82, *SD*=4.18, *t*(26)=3.66, *p=*.001, *d*=0.71, 95%- *CI*=[0.28;1.12]; day 2: *M*=9.33, *SD*=5.35, *t*(26)=4.08, *p*<.001, *d*=0.79, 95%-*CI*=[0.35;1.21]). Notably, no significant differences in task performance could be found for any of the key outcome measures, suggesting that our repeated assessments of motivated behavior were not substantially influenced by undesirable repetition effects. Neither the amount of correct choices (Δ=1.17, *t*(26)=-1.22, *p*=.232), mean confidence ratings (Δ=-0.21, *t*(26)=0.10, *p*=.919), mean effort (Δ=1.21, *t*(26)=-0.60, *p*=.553), mean feedback (Δ=1.51, *t*(26)=-1.45, *p*=.159), nor the coregulation of effort and confidence (Δ=0.07, *t*(26)=-1.24, *p*=.224) differed between examinations. Moreover, confidence, effort, and their coregulation showed moderate to high temporal stability, whereas task success was less stable across a one-week period (see Supplementary Fig. S3). Together, these results indicate that as in experiment 1, subjects learned the probabilistic structure of the task and succeeded to exploit their knowledge of the task by increasing their effort when reward was in reach, but to reduce their effort otherwise (i.e., when the current choice was less likely to be correct). Moreover, the absence of undesirable repetition effects for the parallel versions and overall assessment of test-retest reliability suggests that the task allows a reliable assessment of individual differences for key components of motivated behavior – learning, effort, and their coregulation.

**Figure 3.**
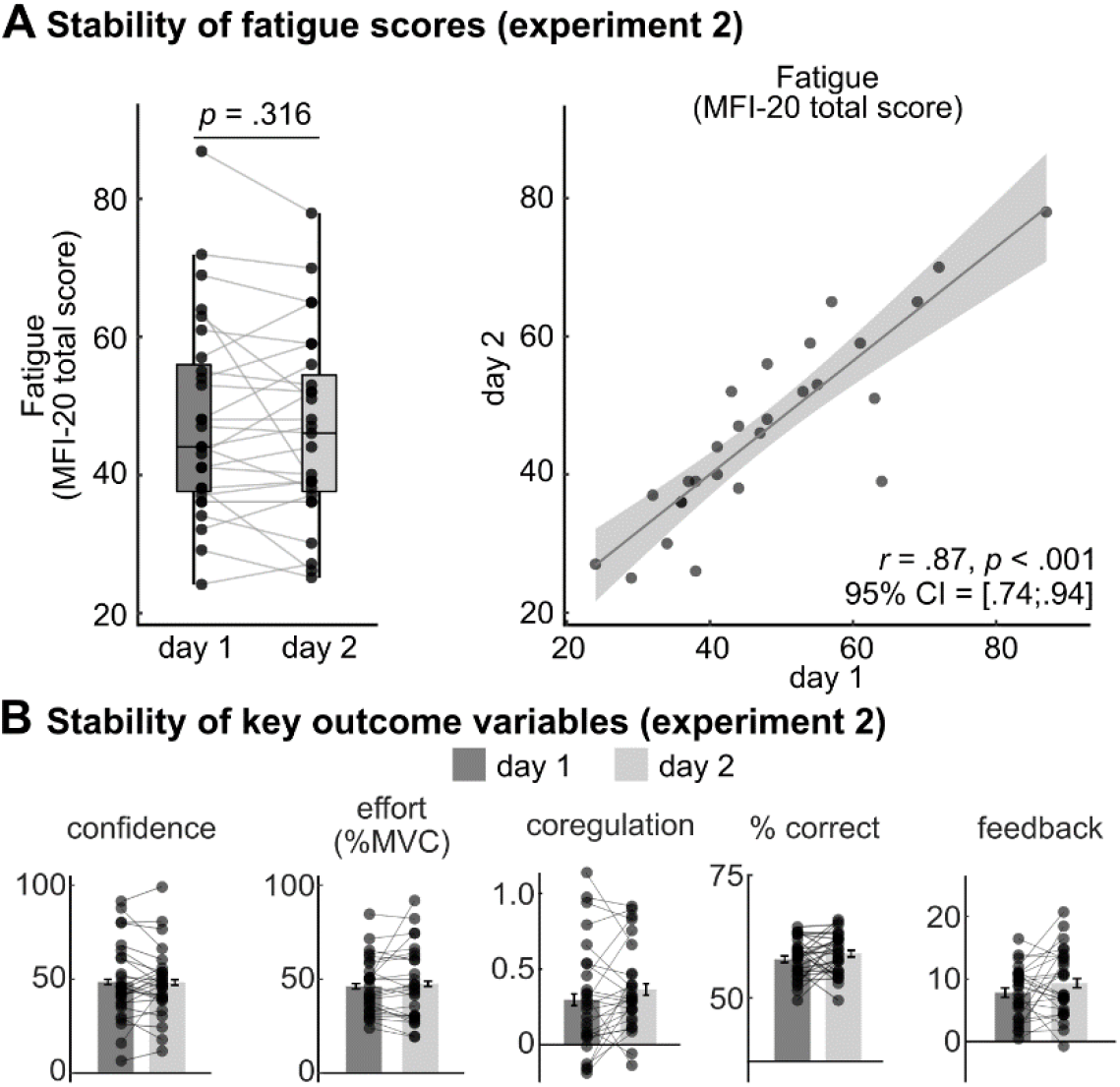
Results from experiment 2. **A** Stability of fatigue measurements using the total score of the 20-item Multidimensional Fatigue Inventory (MFI-20; Smets et al. 1995). **B** Stability of the five outcome variables from the experimental task. None of these variables showed significant changes between test days (see main text).

### Effects of SARS-CoV-2 vaccination-induced state changes of fatigue on motivated behaviour

In experiment 3, subjects (*N*=55) were again invited twice: once following vaccination against SARS-CoV-2 with either AstraZeneca/BioNTech-Pfizer/Moderna (19/25/11) and the other time approximately two weeks before or after vaccination (median time difference of 12.12 days; *IQR*=11.41). MFI-20 scores showed that the vaccination induced state changes in fatigue. On the baseline day, fatigue scores were significantly lower (*mean*=50.13, *SD*=13.70) compared to the measurement after vaccination (*mean*=59.07, *SD*=16.34; Δ=8.95, *t*(54)=4.09, *p*<.001, *d*=.55, 95%- *CI*=[0.26;0.83]; **Fig. 4B**). Fatigue scores were again significantly correlated between days, but the stability of fatigue scores was significantly reduced compared to experiment 2 (*z*=-3.54, *p*<.001). Hence, these data show that controlled, temporally limited immune activation induced state changes in fatigue beyond the stable individual differences at baseline as observed in experiment 2. This is perfectly expected based on the side effects and clinical data on SARS-CoV-2 vaccinations (Anderson et al., 2020; Menni et al., 2021; Polack et al., 2020). We next tested whether individual differences in the experience of fatigue were linked to distinct components of motivated behavior assessed by repeated application of the Learn/Effort-Task.

**Figure 4.**
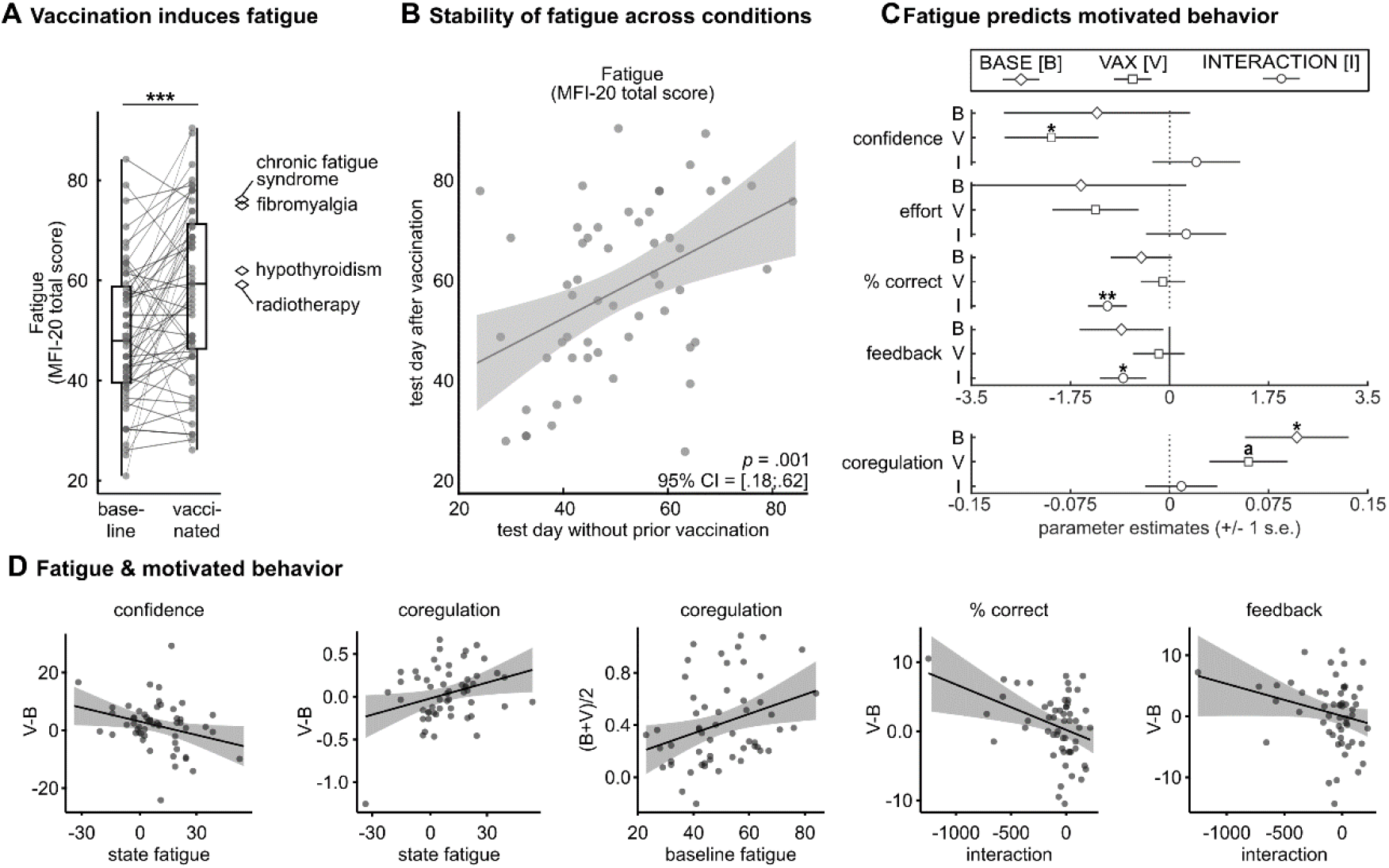
Design and results from experiment 3. **A** Fatigue scores in dependence of vaccination against SARS-CoV-2. Diamonds indicate scores from references groups from other studies (Chronic fatigue syndrome: Natelson et al. (2019), fibromyalgia: Ericsson et al. (2016): hypothyroidism: Gentile et al. (2003); radiotherapy: Lundh Hagelin et al. (2007)). **B** Stability of fatigue scores between days with and without shortly preceding vaccination. **C** Mixed effects regression models were used to test the effects of baseline [B] and vaccination-induced [V] state changes of fatigue and their interaction [I] on measures of motivated behavior. Parameter estimates are shown for B, V, and I, and horizontal lines depict 95% confidence interval (CI) of the regression weight estimates. **D** Scatter plots visualize bivariate associations between predictors and outcome variables without controlling for nuisance regressors. MFI-20 = 20-Item Multidimensional Fatigue inventory (Smets et al., 1995). * = p<.05, ** p <.01.

#### Effects of baseline fatigue

Subjects with higher baseline fatigue showed higher coregulation of confidence and effort (*b*=0.10, *SE*=0.04, *t*(55.00)=2.46, *p*=.017), indicating that in these subjects, effort expenditure was more strongly related to the momentary confidence about their chances to win. However, baseline fatigue scores were not associated with mean effort (comparison to model without fatigue predictors: *X*^2^=0.71, *p*=.400), confidence (*X*^2^=0.61, *p*=.436), nor with any of the two measures of task success (percent correct choices: *X*^2^=0.85, *p*=.357; mean feedback: *X*^2^=1.31, *p*=.253).

#### Effects of state changes in fatigue

Vaccination-induced state changes of fatigue significantly predicted mean confidence during the task (comparison to baseline fatigue model: *X*^2^=6.11, *p*=.013). Specifically, subjects with stronger state changes in fatigue (i.e., larger increases) showed reduced confidence (*b*=-2.09, *SE*=0.83, *t*(65.58)=-2.52, *p*=.014; see **Fig. 4C**). Furthermore, the prediction of coregulation of confidence and effort significantly improved when state changes of fatigue were added as a predictor (*X*^2^=3.88, *p*=.049). However, this was driven by a single influential data point (robust estimate: *b*=0.04, *SE*=0.03, *t*=1.51, 95%-Wald-*CI*=[-0.01; 0.10]; for non-robust estimates see Supplementary Information). State changes in fatigue did not predict effort (comparison to model including baseline fatigue: *X*^2^=2.75, *p*=.097), percentage of correct choice (*X*^2^=0.09, *p*=.766), nor mean feedback (*X*^2^=0.17, *p*=.678).

#### Effects of the interaction of baseline fatigue with state changes in fatigue

Finally, we tested whether motivated behavior was particularly affected if subjects with higher baseline fatigue also experienced greater state changes of fatigue. This reflects the notion of vulnerability-stress models of disease (Ingram & Luxton, 2005) where the interaction of enduring conditions and acute factors like immune-system activation determines subjective burden and behavioral outcomes. Here, the interaction of baseline and state changes of fatigue predicted the percentage correct choices (comparison to state changes of fatigue model: *X*^2^=9.53, *p*=.002). Precisely, subjects with high baseline fatigue and strong responses to the vaccination made more errors (*b*=-0.01, *SE*=0.003, *t*(78.73)=-3.22, *p*=.002). Similarly, the interaction of baseline and state changes of fatigue significantly predicted mean feedback (*b*=-0.82, *SE*=0.41, *t*(72.21)=-2.00, *p*=.049), leading to a marginally significant improvement in model fit (*X*^2^=3.81, *p*=.051). However, baseline and state changes of fatigue did not have interactive effects on effort (*X*^2^=0.17, *p*=.680), confidence (*X*^2^=0.36, *p*=.547), or coregulation (*X*^2^=0.11, *p*=.744). Taken together, these findings illustrate that subjects with already higher levels of baseline fatigue were particularly affected in their task performance in case of strong vaccination-induced state changes of fatigue.

#### Summary of fatigue effect

In sum, our data indicate that variability in various outcome variables from the Learn/Effort-Task can be explained by fatigue and its perturbations after SARS-CoV- 2 vaccination. Specifically, while baseline fatigue predicted increased coregulation of confidence about the effectiveness of own actions and ensuing effort execution, subjects with higher state changes in fatigue experienced reduced confidence during learning. Moreover, subjects with high baseline fatigue and additional strong state changes in fatigue showed diminished task performance in terms of correct choices and effectively received feedback.

## Discussion

Fatigue is a central symptom in various somatic and psychiatric conditions (Chaudhuri & Behan, 2004; Corfield et al., 2016), a hallmark criterion in myalgic encephalomyelitis / chronic fatigue syndrome (ME/CFS) (Afari & Buchwald, 2003), and is associated with “Long-COVID” (Davis et al., 2021; Lidbury & Fisher, 2020; Townsend et al., 2020; World Health Organization, 2021). While such chronic and pathological forms of fatigue cannot be relieved even by extended periods of rest (Karshikoff et al., 2017), rather short-term, state-changes of fatigue are an adaptive reaction of the body to save energy and resolve with behavioral adaptations (Dantzer et al., 2014; Lacourt et al., 2018; Stephan et al., 2016). Here, we took advantage of the ongoing SARS-CoV-2 vaccination campaign to disentangle how vaccination-induced state changes and baseline differences in fatigue affect mental and physical components of behavioral regulation.

As expected, we found that SARS-CoV-2 vaccination induces considerable state increases of fatigue (Anderson et al., 2020; Menni et al., 2021; Polack et al., 2020; Ramasamy et al., 2020). Together with interindividual differences at baseline, these state changes in fatigue predicted determinants of motivated behavior in the task. Precisely, greater state changes of fatigue in the immune activation model were related to reduced confidence during learning. This is in line with studies showing altered learning following thyroid vaccination (Harrison et al., 2016) and with accumulating research indicating detrimental effects of immune parameters on brain systems that support learning (Felger & Treadway, 2017). In contrast, baseline fatigue led to a greater dependency between effort expenditure and confidence, which implies that higher baseline fatigue is related to more targeted investments of physical resources. Thus, greater confidence is needed to express physical effort to obtain rewards. This reflects a greater subjective cost of effort execution, which parallels the observation that increased fatigue also is related to reduced efforts to engage in social interactions (Slavich & Irwin, 2014). Similar findings of increased subjective costs for physical effort are typically observed in clinical samples (Jurgelis et al., 2021; Tran et al., 2021) and have been interpreted as an adaptive bodily response to exhaustion. This is also supported by our finding that state changes of fatigue did not have detrimental consequences in terms of performance outcome. Surprisingly, we did not find evidence for an association of fatigue with average levels of physical effort as previously reported (Jäkel et al., 2021).

Our findings emphasize the relevance of investigating the interplay of mental and physical components of motivated behavior in fatigue to reveal processes that would remain unseen when testing the components in isolation: individually, neither high baseline fatigue, nor strong state changes of fatigue impaired overall task success. Rather, reduced overall task performance only surfaced when vaccination-induced state changes occurred on the background of relatively high baseline fatigue. This was indicated by diminished choice accuracy and lower financial task outcomes. Regardless of the subjective burden of physical activity in fatigue (Holtzman et al., 2019; Lindheimer et al., 2017), this behavioral pattern could suggest a motivational dilemma for individuals with high baseline fatigue and large state increases in fatigue: on the one hand, temporarily constrained inflammation-induced fatigue diminishes the individual’s confidence about when and where effort investments pay off. On the other hand, effortful actions are more strongly guided by momentary confidence in their efficacy. Considered in separation, neither of these fatigue effects is likely to impair overall task success and can be compensated. When combined, however, reduced confidence and a stronger reliance of effort on confidence will lead to less favorable results. Thus, comparable to a vulnerability-stress model (Ingram & Luxton, 2005), people with greater baseline fatigue are particularly prone to the detrimental effects of additional state increases in fatigue (for instance due to an acute immune-response as in the present study), that may then manifest in adverse behavioral outcomes. Such a perspective aligns with the wave-like dynamics in many fatigue-associated clinical conditions, such as in ME/CFS, where short-lived stressful events can exacerbate the disorder (Jammes et al., 2005; Keech et al., 2015; Sorensen et al., 2003). The here described interplay of rather stable differences and temporarily circumscribed responses in the context of immune activation suggests that an exclusive focus on either short-lived peaks or long-lasting, habitual levels of fatigue could hinder the characterization of important mechanisms that may be helpful to understand individual trajectories and effects on everyday life activities.

Notably, in the present study we only had access to a non-clinical setting where behavioral regulation is mostly adaptive and only impacted task performance when greater baseline fatigue was accompanied by vaccination-induced state changes. How this motivational dilemma translates into more pathological forms of fatigue and contributes to the detrimental outcomes of Long-COVID and ME/CFS needs to be considered for a better understanding of these multi-faceted syndromes. In the long run, experimental tasks like ours could thereby help to bridge the gap between studies on fatigue relying on self-report with physiologically informed ideas of how immunological processes contribute to significant behavioral changes via neurobiological pathways (Felger & Treadway, 2017; Müller & Apps, 2019).

## Methods

### Subjects

Across three experiments, a total of 143 subjects were recruited. In experiment 1, two out of 50 subjects were excluded, resulting in a final sample of 48 subjects (mean age=22.31, SD=2.75, 37 female, 11 male), all of whom were university students. One subject was excluded due to low variability of confidence ratings and another due to bad calibration of the hand-dynamometer. In experiment 2, we recruited 30 subjects, all university students or employees from the University of Lübeck and the University Hospital Schleswig-Holstein recruited via university-wide mailing lists; 3 were excluded from data analysis due to insufficient data quality (e.g. no ratings of confidence, force calibration not successful; see below), which resulted in a final sample of 27 subjects (23 female, 4 male; age: mean=32.93 years, *SD*=11.20). The study was a within-subjects design, with two timepoints separated by a median of 7.99 days (*IQR*=7.91). In experiment 3, we recruited 63 subjects, who were university students or employees at the University of Lübeck and the University Hospital Schleswig-Holstein. Subjects were recruited via mailing lists as part of the university-wide vaccination program against SARS-CoV-2 from February to August 2021; 8 were excluded from data analysis due to insufficient data quality (e.g. no ratings of confidence, force calibration not successful; see below), which resulted in a final sample of 55 subjects (42 female, 13 male; age: mean=29.64 years, *SD*=11.09). The study was a within-subjects design, with two experimental groups: members of the first group (Day1Vax, # excluded=5, final *n*=27) were vaccinated one to three days prior to the first experimental day, whereas members of the second group (Day2Vax, # excluded=3, final *n*=28) were vaccinated one to three days prior to the second experimental day (see Table 1). For mRNA vaccines, systemic side-effects of the vaccination including fatigue have been shown to be more frequent and severe following the second dose (Anderson et al., 2020; Menni et al., 2021; Polack et al., 2020), whereas for the AstraZeneca vaccine, the opposite holds (Ramasamy et al., 2020). Therefore, to maximize the expected effects of vaccination on state changes of fatigue, participants being vaccinated with the AstraZeneca vaccine were invited following the first vaccination dose, whereas participants vaccinated with an Mrna vaccine were invited following the second dose. All subjects reported to have normal or corrected-to-normal vision, gave written informed consent prior to participation, and were financially reimbursed following study completion. The study protocol was approved by the local ethics committee of the University of Lübeck (AZ 18-014; AZ 21-136). For safety reasons, and adhering to the university’s safety guidelines, subjects in experiment 2 and 3 were not included in the study if they reported any symptoms of a SARS-CoV-2 infection, were presently quarantined, or had close personal contact with a person infected with SARS-CoV-2 within 14 days prior to participation.

**Table 1.** Characteristics of study groups in experiment 3

|  |  | group |  | group comparison |  |
| --- | --- | --- | --- | --- | --- |
|  |  | Day1Vax | Day2Vax | Test statistic | p |
| female/male | | 19/8 | 5/23 | $\chi^2=1.06$ | .304 |
| age | mean (SD) | 29.19 (10.10) | 30.07 (12.13) | $t(53)=-0.29$ | .770 |
| AZ/ BT/ M | frequency | 16/8/3 | 3/17/8 | $\chi^2=14.39$ | <.001 |
| hours between vaccination and test | median (IQR) | 47.82 (25.34) | 48.95 (21.55) | $U=349$ | .634 |
| days between first and second test | median (IQR) | 12.02 (3.84) | 17.14 (25.35) | $U=309$ | .251 |

**Note.** AZ = AstraZeneca vaccine, BT = BioNTech-Pfizer vaccine, M = Moderna vaccine

### The Learn/Effort-Task

Subjects completed 200 experimental trials on each test day. On each trial, subjects chose between one of two buttons (blue or yellow). After the decision, a flash indicated that subjects could press the hand-dynamometer held in their dominant hand. Subjects were instructed that the physical effort would be rewarded if the correct button had been selected before, but would be penalized otherwise (i.e., points would be subtracted). Subjects won or lost up to 100 points, equivalent to the percentage of their maximum grip force exerted on that trial (henceforth: maximum voluntary contraction, MVC; see Supplemental Material). Thus, if a subject chose the correct button and then exerted 78% of their MVC, 78 points were gained. If, however, the incorrect button had been chosen, 78 points were subtracted (**Fig. 1A**). Therefore, to maximize their points, subjects had to use the information provided by the outcome of each trial to learn when one or the other color was correct, make their choice, and adjust their effort expenditure accordingly. Aside from the binary choice data and the measurements of physical effort, 70 intermittent ratings of subjective confidence regarding the upcoming button choice were acquired. **Fig. 1B** illustrates how both effort and confidence evolved during the task, averaged across all subjects from experiment 1. The input sequence that dictated whether the blue or yellow button was correct on a given trial was predetermined and identical for all subjects. Both colors were correct equally often but the frequency with which one or the other button was correct varied between different phases of the task (black line **Fig. 1B**). On the second day in studies 2 and 3, colors were switched to purple and orange and the left/right locations of buttons on the screen were switched. Physical effort was measured using a hand-dynamometer by Vernier (www.vernier.com, Beaverton, Oregon, USA). Presentation of stimuli was controlled by a computer using Psychtoolbox (Brainard, 1997), a Matlab (Mathworks Inc., Sherborn, MA, USA) based toolbox. In total, the Learn/Effort-Task lasted approximately 45 minutes, excluding instructions and post-experimental interviews. See Supplementary Information for details on the familiarization with and calibration of the hand-dynamometer.

### Psychometric measures

After each session, subjects filled in questionnaires regarding the experiment (e.g. exhaustion, motivation, or overall subjective performance estimates), and other psychometric measures related to the task, including the fatigue during the last days (Smets et al., 1995), handedness (Oldfield, 1971), sleepiness (Johns, 1991), sleep habits (Buysse et al., 1989), sickness behavior (Andreasson et al., 2013), and intolerance of uncertainty (Carleton et al., 2007).

### Statistical Analysis

#### Key outcome measures (experiments 1-3)

We computed five key outcomes measures, including 1) mean confidence ratings, 2) mean effort exertion across the entire task (percentage of MVC, see above), 3) coregulation of confidence and effort (i.e., bivariate Pearson correlation between confidence ratings and % MVC on the following trial; Fisher-z-transformed), and to assess task success, 4) the mean percentage of correct choices for each subject as well as 5) the mean feedback across all trials (i.e., the average number of earned points).

#### Assessment of test-retest reliability (experiment 2)

Test-retest stability of inter-subject differences were computed using Pearson correlation coefficients of the MFI-20 (**Fig. 3A**) as well as of the key outcome variables (**Fig. 3B**) taken on day 1 and day 2.

#### Predicting task behavior with fatigue using mixed effects models (experiment 3)

We used mixed effects models to assess the effects of baseline fatigue, state changes of fatigue (i.e. difference between baseline fatigue score and the fatigue score assessed on the day following SARS-CoV-2 vaccination), and their interaction on task behavior (see **Fig. 4** and Supplementary Fig. S1). Here, mixed effects models allow us to incorporate predictor variables that are stable within persons as well as predictors that vary by test day in a single model, including their joint effects and adjusted for confounding factors. Precisely, we proceeded as follows: in a baseline model, we included only the following predictors of no interest (nuisance regressors): a dummy-coded variable indicating whether a data-point was from the test day with (1) or without (0) prior vaccination; a dummy-coded variable indicating whether it was the first (0) or the second day (1) of participation; the interaction of the two previous predictors; age in years; a dummy-coded predictor indicating each subject’s gender (male=0; female=1). These served to adjust for demographic and test order confounds in the following models: In our first model of interest, we tested for each of the five outcome variables whether baseline fatigue contributed to the model prediction. We defined baseline fatigue as each subject’s fatigue score assessed on the day without prior vaccination. This score was accordingly treated as a subject-specific constant and was entered in the model in addition to the nuisance regressors of the baseline model. In the second model of interest, we tested whether state changes of fatigue induced by prior vaccination explained variability in the outcomes over and above all other predictors, including baseline fatigue. To compute vaccination-induced state changes in fatigue, we subtracted the baseline fatigue score from the fatigue score assessed on the day following SARS-CoV-2 vaccination. Therefore, on the test-day without prior vaccination this variable was always zero, but could take on both positive values (when fatigue was higher following vaccination than at baseline) and negative values (when fatigue was lower following vaccination than at baseline, which was rarely the case; see results). This variable was entered as predictor on top of all nuisance regressors and baseline fatigue. In our final model of interest, we tested the effect of the interaction of baseline and state changes of fatigue as an additional model term, analyzing whether having both high baseline and higher state changes in fatigue could additionally interfere with task performance. We computed the interaction term as the product of group mean-centered baseline fatigue and group mean centered state changes of fatigue on the test day following the vaccination.

All mixed effects models included fixed and random intercepts, as well as fixed slopes for each of the predictor variables. Random slopes were omitted as models were not identifiable with random slopes. All continuous predictors were z-standardized, except for age (which was centered). We used likelihood ratio tests to compare model fits of the first mixed effects model of interest against the baseline model (i.e., testing whether baseline fatigue explained outcome variance), the second model of interest against the first (i.e., testing whether state-changes of fatigue explained outcome variance), and the third model of interest against the second.

### Robustness checks

We estimated robust mixed effects models to safeguard potential effects against outliers and influential data points and computed the Wald confidence intervals for statistical inference of the robust mixed effects models. In case any results from the non-robust and robust analyses were diverging, these discrepancies were described and results of the robust model were reported. In addition, to reassure that our results were not caused by our analytical choice to use mixed effects models, we additionally ran simple linear regression models on difference scores, reproducing all our findings (see Supplementary Table S1).

### Manipulation check

We tested whether subjects used their knowledge of the task to adjust their effort expenditure. If subjects did not adjust their effort expenditure like this, they should show the same level of effort irrespective of whether their choice on a given trial eventually turned out to be correct or not. Thus, we were able to compute an expected mean feedback that would arise through such behavior. Precisely, for each subject we computed the mean feedback that would be expected on correct trials, by multiplying the percentage of correct trials with the average effort shown across all trials. Similarly, we computed the mean feedback that would be expected on incorrect trials by multiplying the percentage of incorrect trials with the average effort across all trials. For instance, for a subject showing an average effort of 65% MVC and choosing correctly on 55% of trials, the expected mean feedback for correct trials would be 65 * .55 = 35.75 points. However, due to 45% incorrect choices, from the expected mean positive feedback we would have to subtract the mean expected negative feedback, i.e.: 65 * .45 = 29.25 points, resulting in an overall expected feedback of 35.75-29.25=6.5 points. Therefore, if the actually received mean feedback was higher than this expected, this would demonstrate that participants did use their expectation of having chosen correctly (or not) on a given trial to adjust their effort expenditure accordingly. Our data confirmed this prediction (see rightmost sub plot in **Fig. 2**).

### Software used for statistical analyses

Data were analyzed using MATLAB R2019b as well as jamovi Version 1.8.2.0 (The jamovi project, 2020) and R (R Core Team, 2018). In R, mixed effects models were computed using the *lme4* package (Bates et al., 2014), robust mixed effects models were computed using the package *robustlmm* (Koller, 2016), and simple linear regression models were run with the inbuilt R-function lm() from the inbuilt R package *stats*. Figures were created using the R package *ggplot2* (Wickham, 2016), the Matlab toolbox *gramm* (Morel, 2018), as well as Inkscape (Inkscape, 2020).

## Data Availability

All data produced and code used in the present study will be available upon reasonable request to the authors.

## Author Contribution

D.S.S., F.L., T.L., F.M.P., and S.K. designed the experiments. D.S.S., F.L., and F.M.P. analyzed the data.

D.S.S., F.L., F.M.P., and S.K drafted the report, all authors discussed the results and revised the report.

## Acknowledgment

We thank Julie Lasselin for providing extensive comments on this manuscript.

## Conflict of Interest

The authors report no conflict of interest.

## Funding

This study received funding from the COVID-19 Research Initiative Schleswig-Holstein.

## Notes

### Competing Interest Statement

The authors have declared no competing interest.

### Author Declarations

The Ethics committee of the University of Lübeck gave ethical approval for this work.

