## Supplements for "Determinants of motivated behavior are linked to fatigue and its perturbation by SARS-CoV-2 vaccination"

### Supplementary Methods

#### Familiarization with the hand-dynamometer

Before reading the instructions for the Learn/Effort-Task, subjects had the opportunity to familiarize themselves with the hand-dynamometer. To this end, the image of a flash was presented centrally on the screen for ten seconds and subjects were instructed to press the hand-dynamometer with varying force to observe how the size of the flash continuously changed in close dependence of their applied force.

#### Calibration of MVC

Following familiarization, subjects first completed a task that was intended to motivate them to exert effort with their maximum voluntary contraction (MVC). On 15 trials, subjects were shown the outline of the flash they had already seen during the familiarization period. They were instructed that they should now try to collect as many stars as possible by pressing the hand-dynamometer. Precisely, exerting effort would increase the size of a white flash image, and the goal on each trial was to exert sufficient effort so that the target outline would be filled in completely by the white flash image. On trials in which the target was reached (i.e., the outline was filled completely), the image of a star popped up on the screen, but if the target was not reached, the image of a star crossed by a red X was presented.

Critically, the size of the target outline changed from trial to trial in dependence of the participant's behavior. On successful trials, the outline's size increased by a factor of 1.075 with a probability of 67%, and otherwise decreased by a factor of .925. On unsuccessful trials, the outline's size decreased by a factor of .925 with a probability of 67%, and otherwise increased by a factor of 1.075. Therefore, the effort needed to reach the target tended to increase from trial to trial, so that we could iteratively approximate each participant's MVC. The maximum force applied during the familiarization period was used to determine the initial scaling of the target outline on the first calibration trial.

MVC was determined as the maximum force applied at any point during the familiarization period or the 15 calibration trials.

The reasons for utilizing this gamified procedure were twofold: First, we did not want participants to guess that their payoff in the experiment depended on their MVC and thus voluntarily show a reduced level of effort during this calibration phase in order to gain points more easily later on. Second, there is extensive literature on the motivational potential of gamification (Buckley and Doyle 2016; Sailer et al. 2013), which could therefore help leading participants to exert high levels of effort during calibration.

**Supplementary analyses for simple linear regressions**

To test the robustness of our findings across a variety of analytical approaches, in experiment 3 we additionally ran simple linear regression models to test the effects of baseline fatigue, state fatigue, and their interaction to the five outcome variables. In contrast to the mixed effects models, only one value for each variable (i.e., regressors as well as dependent variables) could be used in these analyses. Next, we describe how the dependent variables were computed, and how this relates to the logic of the mixed effects models.

Baseline fatigue for each participant was defined to be the fatigue score reported on the test day without prior vaccination. In the mixed models, this value was equal on both days for each participant, and thus predicted the average value of a given outcome variable (e.g., % correct choices) across both days. Accordingly, the average of a given outcome variable across both days for each participant was used as the dependent variable when testing the effect of baseline fatigue using simple linear regression. As nuisance regressors, we included dummy variables (including only 0s or 1s) coding group and gender, as well as a continuous predictor to account for variability linked to age.

State fatigue for each participant was defined as the change in fatigue from the test day *without* previous vaccination to the test day *with* previous vaccination (which was significantly positive; see main text). Thus, in the mixed models, the fixed slope of state fatigue estimated the association between vaccination-induced changes in fatigue and vaccination-induced changes in a given outcome variable. Therefore, we computed change scores for both fatigue and the given outcome variable (test day with previous vaccination minus test day without previous vaccination) that were used as regressor and dependent variable, respectively. In addition, we controlled for the effects of baseline fatigue, study group, gender, and age.

Last, in the regression models testing the interaction of baseline and state fatigue, the outcome variables were computed in the same way as in the models for state fatigue (i.e., difference scores). The interaction term was computed as the product of baseline fatigue and state fatigue. Since state fatigue was 0 for all subjects on the day without previous vaccination, so was the interaction term, and only the day with previous vaccination was analyzed. Here, age, gender, study group, baseline fatigue, and state fatigue were included as nuisance regressors.

In these analyses, all continuous predictors and dependent variables were z-transformed.

### Supplementary results

#### Experiment 1

A total of 50 participants (mean age=22.24 years, SD=2.72, 38 female, 12 male) were recruited at the University of Lübeck to participate in the pilot study. All participants gave written informed consent prior to participation and received a monetary compensation or partial course credit for taking part in the study. Two subjects were excluded from further analysis: one due to low variability of confidence ratings and another due to bad calibration of the hand-dynamometer. Hence, data for a total of 48 participants were analyzed (mean age=22.31, SD=2.75, 37 female, 11 male).

We used this dataset to characterize behavior in the Learn/Effort-Task for the first time. Across all 70 ratings, subjects reported a mean confidence of 40.51 (SD=13.75), and showed a mean effort of 45.21 (% maximum voluntary contraction, MVC; SD=11.52). The mean coregulation of confidence and subsequent effort, i.e., their Fisher-transformed Pearson correlation, was .45 (SD=.30), and differed significantly from 0 ($t(47)=10.58$, $p<-001$, $d=1.53$, 95% CI=[1.11;1.94]). This demonstrates that the amount of effort expended was robustly related to the subjective confidence that it would entail a reward.

On average, participants selected the correct button on 59.12% of trials (SD=3.48), which was significantly above chance level ($t(47)=18.15$, $p<.001$, $d=2.62$, 95% CI=[2.02;3.22]). This shows that participants succeeded to learn about the probabilistic structure of the task. Finally, across all trials, participants received an average of 9.73 points (SD=4.41), which was significantly more than 0 ($t(47)=15.30$, $p<.001$, $d=2.21$, 95% CI=[1.68;2.73]). Moreover, for each participant we computed the expected feedback under the assumption that the level of effort expended did not differ between trials with correct and incorrect choices. To do so, we computed the mean expected positive feedback and subtracted thereof the mean expected negative feedback (e.g., correct choices on 55% of trials and mean effort = 65% MVC: $(.55 * 65) – (.45 * 65) = 6.5$). A paired t-test showed that the true mean feedback was significantly higher than this expected feedback ($t(47)=8.67$, $p<.001$, $d=1.25$, 95% CI=[0.87;1.63]). This demonstrates that participants successfully exploited their knowledge of the task's probabilistic structure by exerting effort when it was likely to be rewarded and by avoiding to do so otherwise (i.e., when expecting a subtraction of points).

Together, these data demonstrate the utility of the Learn/Effort-task for assessing, simultaneously, how individuals learn whether their efforts will yield rewarding outcomes and how they use the information acquired during learning for the adjustment of their physical effort.

**Experiment 2**

In a post- experimental manipulation check, participants estimated to have chosen correctly on 57.78% of the trials (*SD* =13.89%) on day 1 and 54.81% (*SD* =13.51) on day 2. Interestingly, this value was only significantly different from a chance performance of 50% on day 1 ($t$ (26)=2.91, $p$ =.007, $d$ =0.56, 95%-*CI* =[0.15; 0.96]) but not on day 2 ($t$ (26)=1.85, $p$ =.076, $d$ =0.36, 95%-*CI* =[0.04; 0.74]). However, both were not significantly different from each other (difference: Δ=- 2.96, $t$ (26)=1.03, $p$ =.311, $d$ =-0.20, 95%-*CI* =[-0.58; 0.18]). Subjectively rated choice performance was not significantly lower than objectively measured (day 1: $t$ (26)=0.05, $p$ =.962, $d$ =0.01, 95%-*CI* =[- 0.37; 0.39]; day 2: $t$ (26)=1.73, $p$ =.095, $d$ =0.33, 95%-*CI* =[- 0.06; 0.72]), indicating that participants did not massively underestimate their own performance.

**Experiment 3**

**Non-robust prediction of coregulation with state fatigue**

Non-robust mixed model analyses suggested a significant prediction of coregulation by state fatigue ($b$=0.06, *SE*=0.03, $t$(79.17)=2.02, $p$=.046).

**Supplementary Figures**

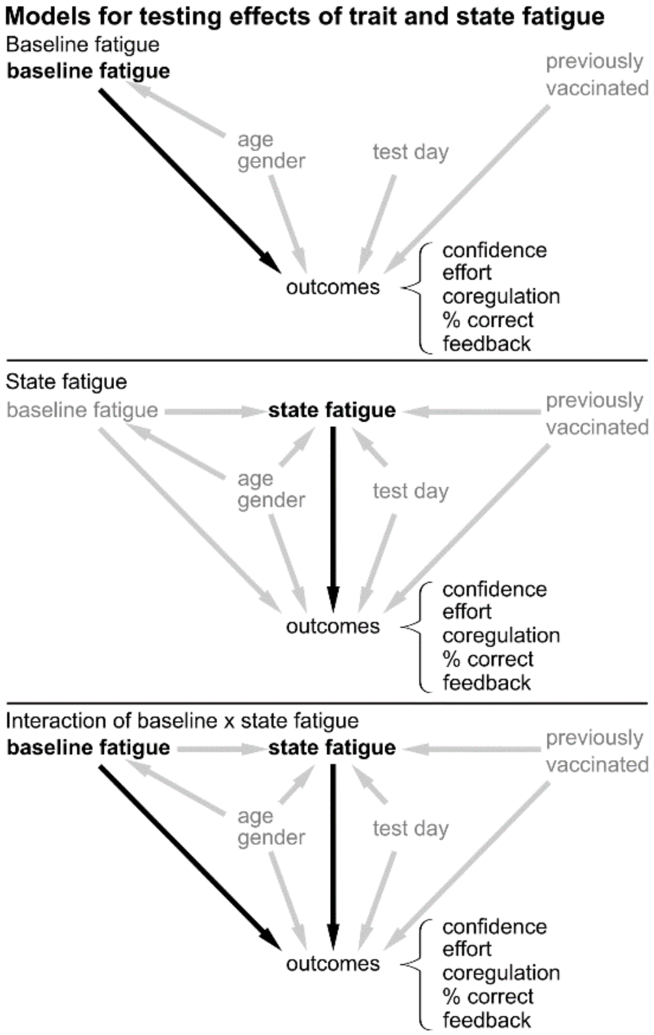


**Supplementary Figure S1**. Directed Acyclic Graphs (DAGs) depicting the rationale for running mixed effects regression models used to test the effects of fatigue on objective measures of motivated behavior. Nuisance regressors in each model are shown in grey and regressors of interest in each model are shown in black and boldface. For each of the five outcome variables, we first estimated the model for baseline fatigue, and then sequentially proceeded to the models for state fatigue and the interaction of baseline fatigue and state fatigue. This approach was identical for both non-robust and robust model estimations.

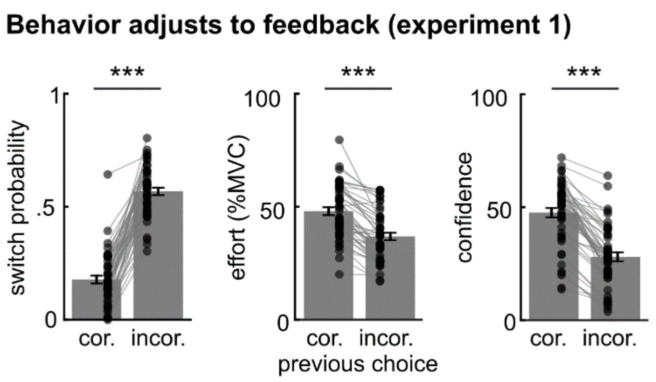


**Supplementary Figure S2**. Results from experiment 1. Effects of preceding choice (correct vs. incorrect) on probability to switch choice (left), effort (middle), and confidence ratings.
MVC = maximum voluntary contraction.
*** = p<.001.

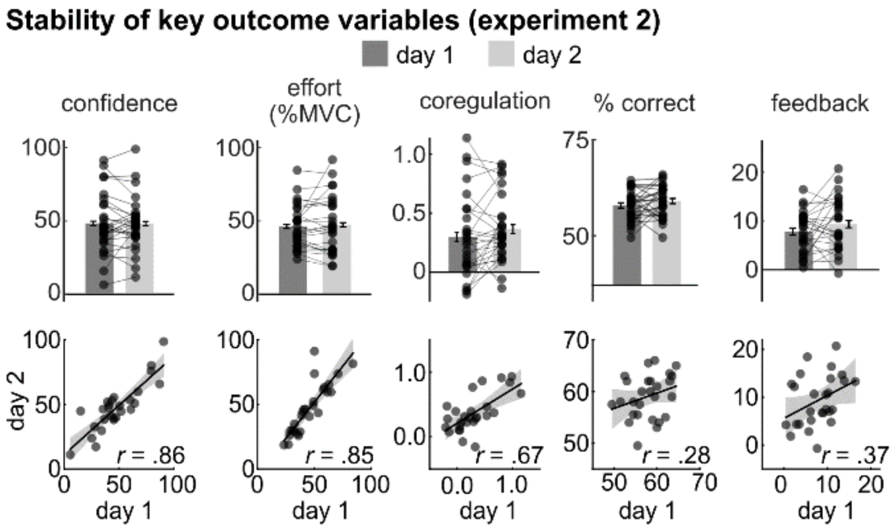


**Supplementary Figure S3**. Results from experiment 2. **Top row** Results as described and depicted in the main manuscript. **Bottom row** Scatter plots show values of outcome variables in the top row plotted for study day 2 (y-axis) against day 1 (x-axis).
MVC = maximum voluntary contraction.
*r* = Pearson correlation coefficient.

### Supplementary Tables

**Supplementary Table S1.** Results from simple regression models

| | baseline fatigue | | | |
|---|---|---|---|---|
| Outcome variable | *B* | *SE* | *t* | *p* |
| confidence | -0.10 | 0.13 | -0.75 | .460 |
| mean effort | -0.11 | 0.13 | -0.80 | .425 |
| coregulation | 0.30 | 0.13 | 2.35 | **.023** |
| % correct choices | -0.11 | 0.13 | -0.88 | .382 |
| mean feedback | -0.15 | 0.14 | -1.10 | .278 |
| | state fatigue | | | |
| | *B* | *SE* | *t* | *p* |
| confidence | -0.37 | 0.15 | -2.47 | **.017** |
| mean effort | -0.29 | 0.14 | -2.07 | **.044** |
| coregulation | 0.38 | 0.15 | 2.50 | **.016** |
| % correct choices | -0.04 | 0.14 | -0.32 | .750 |
| mean feedback | -0.09 | 0.13 | -0.67 | .506 |
| | interaction baseline x state fatigue | | | |
| | *B* | *SE* | *t* | *p* |
| confidence | -0.02 | 0.41 | -0.06 | .956 |
| mean effort | 0.10 | 0.38 | 0.25 | .801 |
| coregulation | 0.29 | 0.41 | 0.71 | .482 |
| % correct choices | -1.09 | 0.35 | -3.07 | **.003** |
| mean feedback | -0.73 | 0.36 | -2.06 | **.045** |

**Note.** Additional nuisance regressors in each model were age, gender, study group, baseline fatigue (for state model and interaction model), and state fatigue (for interaction model).
